# Temporal contact patterns and the implications for predicting superspreaders and planning of targeted outbreak control

**DOI:** 10.1101/2023.11.22.23298919

**Authors:** Rachael Pung, Josh A Firth, Timothy Russell, Tim Rogers, Vernon J Lee, Adam J Kucharski

## Abstract

Epidemic models often heavily simplify the dynamics of human-to-human contacts, but the resulting bias in outbreak dynamics – and hence requirements for control measures – remains unclear. Even if high-resolution temporal contact data were routinely used for modelling, the role of this temporal network structure towards outbreak control is not well characterised. We address this by assessing dynamic networks across varied social settings in three ways. Firstly, we characterised the distribution of retained contacts over consecutive timesteps by developing a novel metric, the “retention index”, which accounts for the change in the number of contacts over consecutive timesteps on a normalised scale with the extremes representing fully static and fully dynamic networks. Secondly, we described the repetition of contacts over the days by estimating the frequency of contact pairs occurring over the study duration. Thirdly, we distinguish the difference between ‘superspreader’ and infectious individuals driving ‘superspreading events’ by estimating the connectivity of an individual (i.e. individual has high connectivity in a timestep if he accounts for 80% of the contacts in the timestep) and the frequency of exhibiting high connectivity. Using 11 networks from 5 settings studied over 3–10 days, we estimated that more than 80% of the individuals in most settings were highly connected for only short periods. This suggests a challenge to identify ‘superspreaders’, and more individuals would need to be targeted as part of outbreak interventions to achieve the same reduction in transmission as predicted from a static network. Taking into account repeated contacts over multiple days, we estimated simple resource planning models might overestimate the number of contacts made by an infector by 20%–70%. In workplaces and schools, contacts in the same department accounted for most of the retained contacts. Hence, outbreak control measures would be better off targeting specific sub-populations in these settings to reduce transmission. In contrast, no obvious type of contact dominated the retained contacts in hospitals, so reducing the risk of disease introduction is critical to avoid disrupting the interdependent work functions. This study identified key epidemiological properties of temporal networks that potentially shape outbreak dynamics and illustrated the need for incorporating such properties in outbreak simulations.

**Significance:** Directly transmitted infectious diseases spread through social contacts that can change over time. Modelling studies have largely focused on simplifying these contact patterns to predict outbreaks but the assumptions on contact patterns may bias results and, in turn, conclusions on the effectiveness of control measures. An ongoing challenge is, therefore, how to measure key properties of complex and dynamic networks to facilitate the development of network disease simulation models, which ensures that outbreak analysis is transparent and interpretable in the real-world context. To address this challenge, we analysed 11 networks from 5 different settings and developed new metrics to capture crucial epidemiological features of these networks. We showed that there is an inherent difficulty in identifying individual ‘superspreaders’ reliably in most networks. In addition, the key types of individuals driving transmission vary across settings, thus requiring different outbreak control measures to reduce disease transmission or the risk of introduction. Simple models to mimic disease transmission in temporal networks may not capture the repeated contacts over the days, and hence could incorrectly estimate the resources required for outbreak control. Our study characterised temporal network data in epidemiologically relevant ways and is a step towards developing simplified contact networks to capture real-world contact patterns for future outbreak simulation studies.

## Background

Directly transmitted infections spread through human social contacts, but the dynamic and often high-dimensional nature of these networks has historically made them difficult to measure and interpret. As a result, epidemic models often implicitly approximate complex dynamic networks with simpler contact processes, including static networks (1, 2), branching processes (3) and compartmental models (4). These relatively simpler models of disease transmission have been well-studied, but it remains unclear how they compare with real-life temporal social networks, which exhibit a mix of repeated and occasional contacts (5, 6). As such, the assumptions in these simpler models could bias model outputs that are crucial for epidemic planning and response, from estimating the required resources for contact tracing and testing programmes to assessing the impact of social distancing measures and vaccine coverage (7–9).

There has been recent progress in the collection of dynamic contact network data via proximity sensors (10, 11) or mobile devices (12). The automated nature of such data collection enabled large-scale deployment for contact tracing during the COVID-19 pandemic (11, 13). These devices work by exchanging radio frequency identification (RFID) signals within a calibrated distance, enabling us to monitor contacts and map the emerging network structure. This can – in theory – enable us to interpret the transmission process on temporal networks. However, in practice most studies still tend to simplify the temporal network structure by extending static network properties, which depend on the characteristics such as population sizes (5), making it hard to compare findings across studies. Furthermore, it can be challenging to tease out the effects of different network features on the transmission dynamics in models (5, 14, 15). Finally, temporal contact data in some studies was collected through self-reported contact dairies, which may be prone to recall bias (6, 14, 16). With the extensive data collected from automated devices, this is increasingly an opportunity to better compare contact structures and hence, the implications for key transmission processes.

Using real-world temporal social data from over 4 million contact events collected across five settings (cruises, community, schools, hospital and workplaces), we quantified the impact of dynamic contacts on key epidemiological metrics driving person-to-person transmission across these varied social settings. As well as examining the range of bias introduced by common simplifying assumptions, we identify the extent to which it is possible to identify individuals linked to superspreading events reliably. To characterise time-varying properties of the real-life networks, we developed a new metric – the retention index – that allows complex dynamic networks to be summarised and compared in an epidemiologically meaningful manner.

## Methods

### Temporal contact network data

We collated temporal contact network data from previously published studies across different settings, with contacts recorded using proximity sensors or mobile devices (Table 1). These devices were calibrated to record contacts between pairs of individuals within a specified radius on cruises and in a community or, alternatively face-to-face interactions in high schools, a hospital and workplaces. The radius approach is omnidirectional, while the face-to-face methods record a contact when the sensors face each other. For each network, we performed preliminary analysis to identify common types of contact, contact durations, and delays before the next contact occurs between a pair of individuals (Table 1). Contact data from the cruises were recorded in 15-second intervals, while in all other networks, contacts were recorded in 5-minute- or 20-second intervals.

**Table 1.** Characteristics of real-world contact networks.

| Network setting | Study date, observed days | Types of contact | Median contact duration (sec) | Median delay in contact (sec) | Remarks (references) |
| --- | --- | --- | --- | --- | --- |
| Cruises, Singapore | Nov 2020, 3d<br>Nov 2020, 3d<br>Jan 2021, 3d<br>Feb 2021, 3d<br>(i.e. four sailings with two in Nov 2020) | P: passenger<br>C: crew<br>P-P (same cabin)<br>P-P (different cabin)<br>C-C (same department)<br>C-C (different department)<br>P-C | 900 for all four sailings | 900 for all four sailings | COVID-19 restrictions onboard. Undirected network (11) |
| Community, Haslemere, UK | Oct 2017, 3d | Household<br>Non-household | 300 | 600 | No data before 0700 hrs and after 2300 hrs. Directed network (12) |
| High Schools, Marseilles, France | Dec 2011, 4d<br>Nov 2012, 7d<br>Dec 2013, 5d | Classmates<br>Non-classmates | 20 for all three high school | 140<br>120<br>100 | No data over weekends. Directed network (16, 17) |
| Hospital, Lyon, France | Dec 2010, 5d | Same department<br>Different department | 20 | 140 | Directed network (18) |
| Workplaces, France | Jun 2013, 10d<br>2015, 10d | Same department<br>Different department | 20 for both workplaces | 220<br>120 | No data over weekends. Directed network (19, 20) |

To analyse the network properties, we first needed to choose a timescale for defining a ‘contact’ within each dataset. In our main analysis, we set the length of the timestep for each network based on the median delay in contact. The timestep was set at 15-min, or 1-hr for subsequent sensitivity analysis. We also performed additional sensitivity analysis, assuming the directed contact networks in the non-cruise settings were undirected. For the high school, hospital and workplace networks, a small timestep (e.g. 20-sec) would result in few repeated contacts over consecutive timesteps because the median delay between contact events was higher than the contact duration (Table 1). As such, the main analysis considered the contact patterns based on timesteps defined for each network, while our sensitivity analysis standardised the timesteps across all networks. A contact is defined to occur within a timestep if it lasts for at least the median contact duration for respective networks (Table 1). At one theoretical extreme, networks may exhibit no variation over time, resulting in a static network, where the contacts remain the same over consecutive time steps; at the other extreme, we have fully dynamic networks, where every individual’s contacts are drawn randomly at each time steps (Figure 1). When simulating the fully dynamic network across consecutive time steps, we retained the degree distribution of each individual observed in a time step but randomly rewired their contacts. This ensures that the fully dynamic network has the same degree distribution as the static network of the same time step.

**Figure 1.**
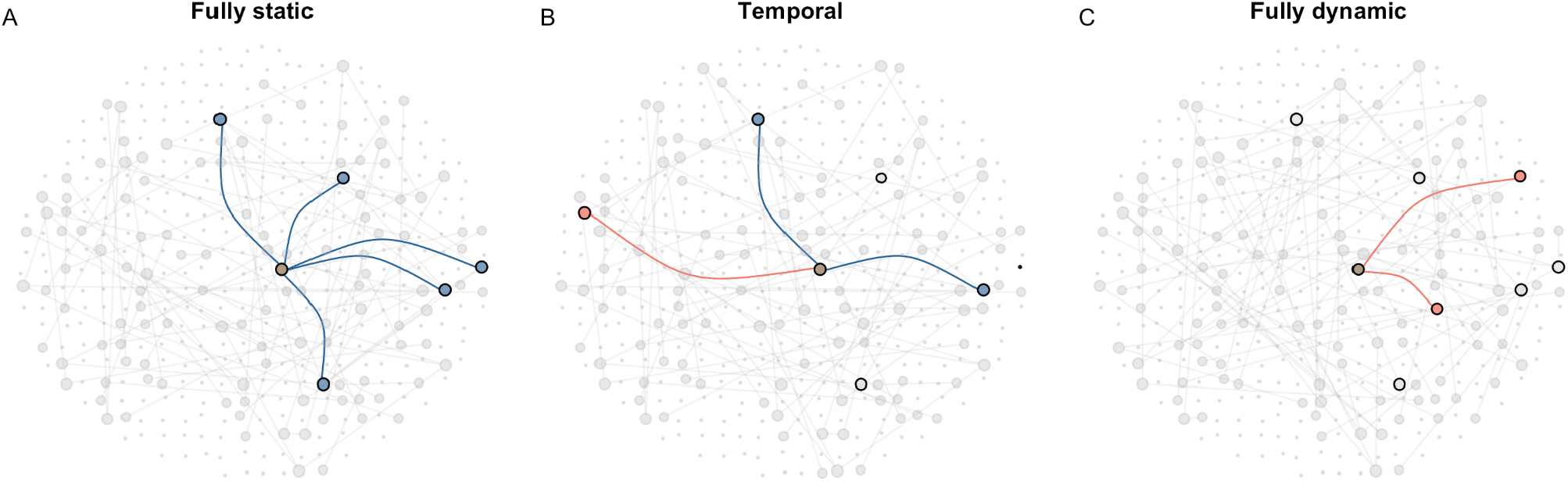
Contacts made by an individual of interest (brown, centre) in a single time step with contacts retained from the previous time step (blue), contacts that were not retained from the previous time step (grey with black outline) and new contacts in current time step (red) for (A) fully static; (B) temporal; and (C) fully dynamic network.

### Contact retention

To explore how contacts were retained and changed over time, we defined the distribution of the number of retained contacts, *r*, over consecutive time steps, *t* and *t +* 1, in the network as follows:

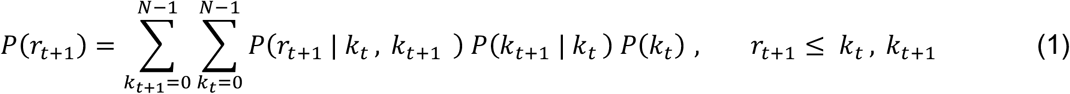

where *k*_*t*_ is the number of contacts (i.e. degree) in time step *t* and *N* is the number of individuals in a network. The maximum possible number of contacts an individual could make is *N*-1. For static or fully dynamic networks, where contacts are either fixed or made at random, *P*(*r*_*t*+1_ | *k*_*t*_, *k*_*t*+1_) of equation (1) is replaced with the binomial distribution as follows:

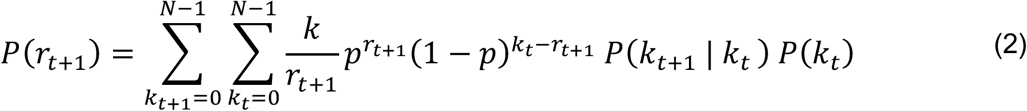

where *k* is the minimum of *k*_*t*_ and *k*_*t*+1_ and *p* is the binomial probability of preserving a contact between a pair of individuals. For static networks, *p* = 1 and equation (2) simplifies as follows

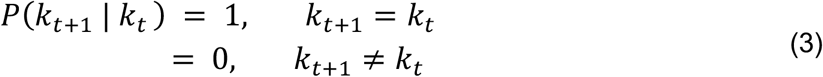

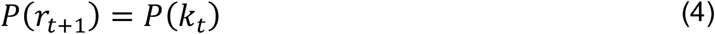

For fully dynamic networks with randomly made links, 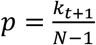 and equation (2) is expressed as follows

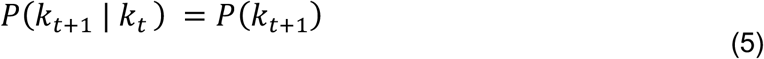

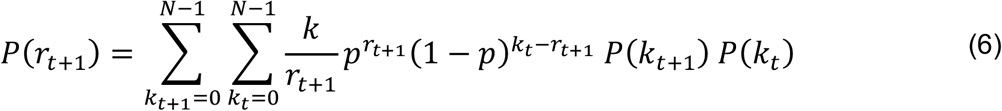

By definition, we expect the highest mean number of retained contacts to be observed in static networks, 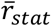, and the lowest in fully dynamic networks,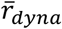. To quantify the mean number of retained contacts in our collated temporal networks, 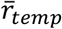, we computed a scaled metric, defined as the ‘retention index’, as follows

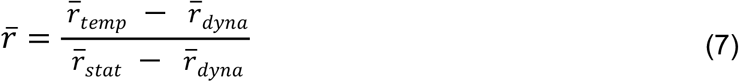

This metric (retention index) provides a standardised measure of where a network lies between the two theoretical extremes. If 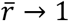, the temporal network reflects a fully static (and hence fully predictable) structure; when 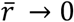, the temporal network reflects a fully dynamic (and hence non-predictable) structure.

### Epidemiological metrics

If contacts are retained over consecutive time steps, it will result in a longer duration of continuous contact and, hence, a higher risk of transmission. Under the assumption that infection does not change the individual’s contact patterns (e.g. for an infection that exhibits substantial asymptomatic or pre-symptomatic transmission), clustering of retained contacts would also limit further disease transmission by an infector if the contact is already infected. To identify predictors of contact retention over consecutive time steps, we estimated the proportion of repeated contacts occurring for each type of contact (Table 1). Besides evaluating the retention of contacts over consecutive time steps, we can also evaluate the repetition of contacts over different days by estimating the frequency distribution of contact encounters in days among all the contact pairs.

We also assessed the bias introduced when assuming independence of contacts over the days. To do this, we estimated the difference between the cumulative unique contacts from the start to the day of interest, and the sum of unique contacts each day from the start to the day of interest. We estimated the relative difference in contacts to generalise the findings across different studies with different population sizes.

### Extent of superspreaders and superspreading events

We defined potential ‘superspreaders’ as individuals frequently identified to account for the top 80% of the contacts made or contact duration over the observed period (see example in next paragraph). We also define potential ‘superspreading events’ to be transmission driven by individuals less frequently identified to account for the top 80% of the contacts or contact duration over the observed period. The latter group of individuals typically forms few contacts. However, for a small proportion of the time, they have many or prolonged contacts and could disproportionately account for many transmission events in that time if they were infectious (21, 22).

In each time step, we identified the individuals accounting for the top 80% of contacts or contact duration (i.e. highly connected individuals). The minimum and maximum proportion of time steps that an individual was identified in this top group could range between 0 to 1. For each incremental proportion of time, we estimated the proportion of the population identified for the corresponding time. To illustrate the extent of transmission events driven by superspreader or superspreading dynamics, we plot the cumulative proportion of the population identified for at least a given proportion of time. For example, we might identify a certain proportion of the population to be highly connected in at least half of the number of observed time steps. In this example, we could label this group as ‘superspreaders’. On the other hand, we might identify a certain proportion of the population to be highly connected but only in less than half of the number of observed time steps. We could label this group as individuals who drive ‘superspreading events’.

To provide context of how the real-world networks compare with static and fully dynamic networks when visualising our results, we simulated a homogenous and an overdispersed network over different time steps to estimate the above metrics. In a homogenous network, expected 80% of the population accounts for 80% of the contacts (i.e. *p*_80_ = 0.8), while in an overdispersed network, this is less than 80% of the population (in this study, we used 50%, i.e. *p*_80_ = 0.5). For a static network, regardless of a homogeneous or an overdispersed network, the same proportion of the population was identified across all time steps by definition. For a fully dynamic network of varying time steps, the proportion of the population identified for each incremental proportion of time is approximately *p*_80_ raised to the power of *s*, where *s* is the number of time steps corresponding to the proportion of time.

## Results

### Contact retention

We found considerable variation in the retention index 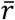 across different networks and over time. For example, cruise networks exhibited an 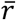 of 0.59 (IQR 0.52–0.81). This study was conducted under strict COVID-19 physical distancing and social gathering restrictions onboard the cruises (Figure 2A). As a result, most of the repeated contacts occurred among passengers who shared the same cabin and hence were in the same travelling group, and crew members of the same department (Figure 1B). We estimated an 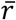 of less than 0.5 in only 12–24% of the observed time steps for the four cruise sailings, indicating that in a given time period, contacts are much more likely to be retained rather than new contacts being made. Between 30–60% of these time steps with lower 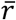 occurred between 1200-1400 hrs and 1800-2000 hrs across the four cruise sailings. Passengers were likely to be engaged in dining during these periods and previous work showed that dining settings promote social contact (11). The seating arrangements or the movement patterns (e.g. buffet counters) facilitate increased mixing and interaction between passengers of different cabins (Figure 2B and Supplementary Figure 1). High values of 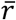 were also observed at the start and end of each day, the result of contact between passengers from the same cabin.

**Figure 2.**
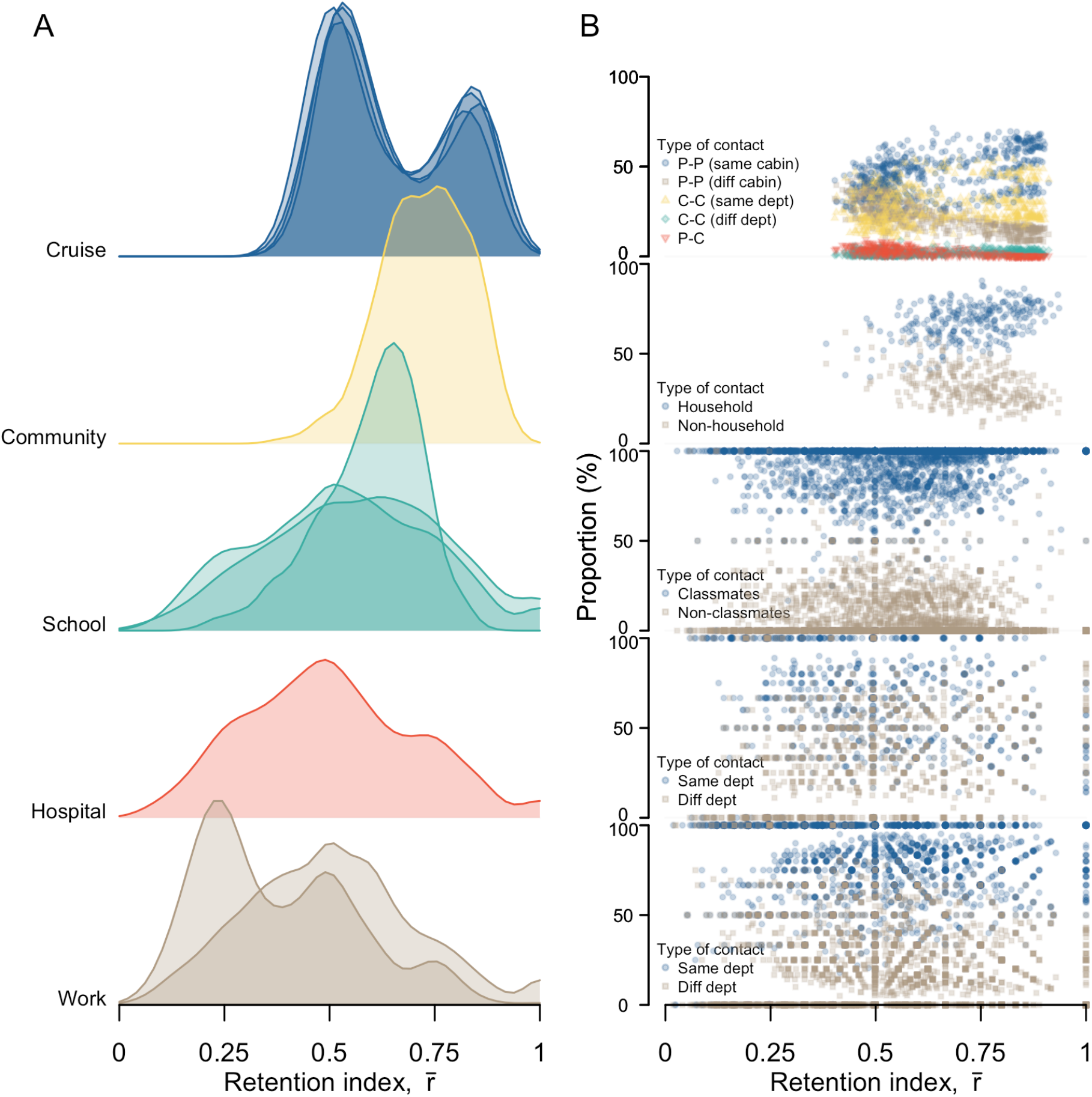
Contacts patterns in different network settings, (a) ridgeline plot showing distribution of contact retention index, 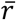, over consecutive timesteps, (b) proportion of the type of retained contacts for respective 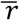.

Pre-pandemic community networks from the UK exhibited an even higher 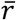 f 0.73 (IQR 0.65– 0.81). About 40% of the contacts occurred before 0900 hrs and after 1700 hrs when the individual is likely to be at home with household contacts (Figure 2A and B, and Supplementary Figure 1). In contrast, networks from schools, a hospital and workplaces showed lower 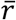 of 0.58 (IQR 0.44– 0.69), 0.49 (IQR 0.36–0.64) and 0.50 (IQR 0.33–0.61) respectively. In these networks, 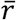 was below 0.5 for about half of the observed duration and changes in 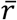 did not exhibit any time trends, unlike the cruise or community networks (Supplementary Figure 1). Moreover, at low and at high values of 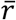, there was no apparent variation in the type of retained contacts. We estimated that contacts made between classmates or individuals of the same department form the majority of the contacts in each time step for the high school network, about 60% for the hospital network and about 80% for the workplace networks. We observed similar proportions among the retained contacts (Figure 2B).

The overall patterns in our analysis remained unchanged when we performed sensitivity analyses around choice of time step and contact definition. We obtained similar results when assuming undirected contacts in the non-cruise settings (Supplementary Figure 2), although when using fixed time steps of 15-min or 1-hr for all networks, the overall median *r* of all networks was slightly lower than the main analysis. However, 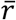 in both the cruise and community networks remained higher than networks from schools, a hospital and workplaces (Supplementary Figure 3 and 4).

**Figure 3.**
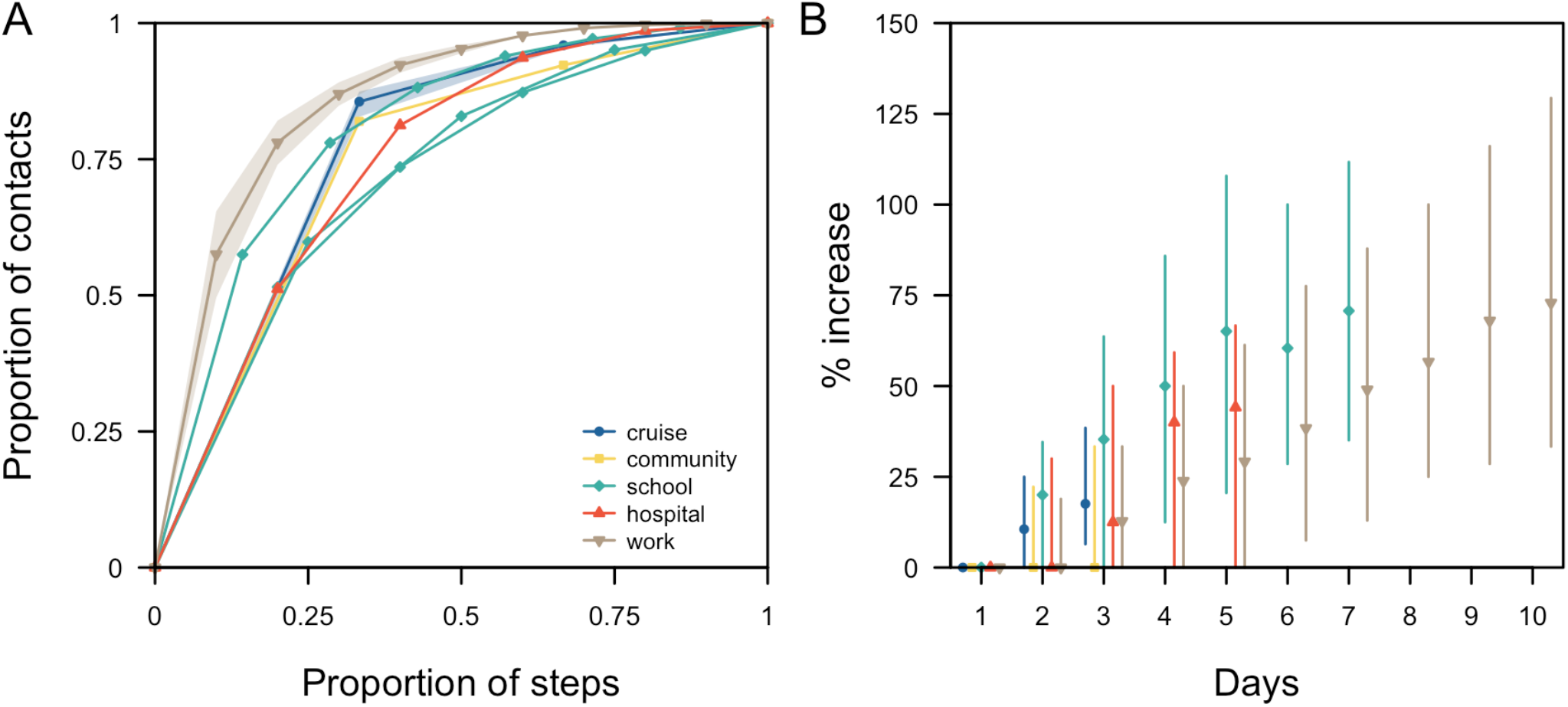
Contact pairs over the study duration in different networks, (A) cumulative distribution of contact encounters in days in pairs of contact. Study duration varied across networks and was normalised. For networks with the same study duration, such as the four cruises and three workplace networks, the distribution was represented by the median (lines) and range (shaded region). For networks with different study durations, such as the three high school networks, or a single network study, such as the community and hospital networks, the distribution of each network study was illustrated, (B) Median (shapes) and range (lines) of the relative difference in the number of unique contacts.

**Figure 4.**
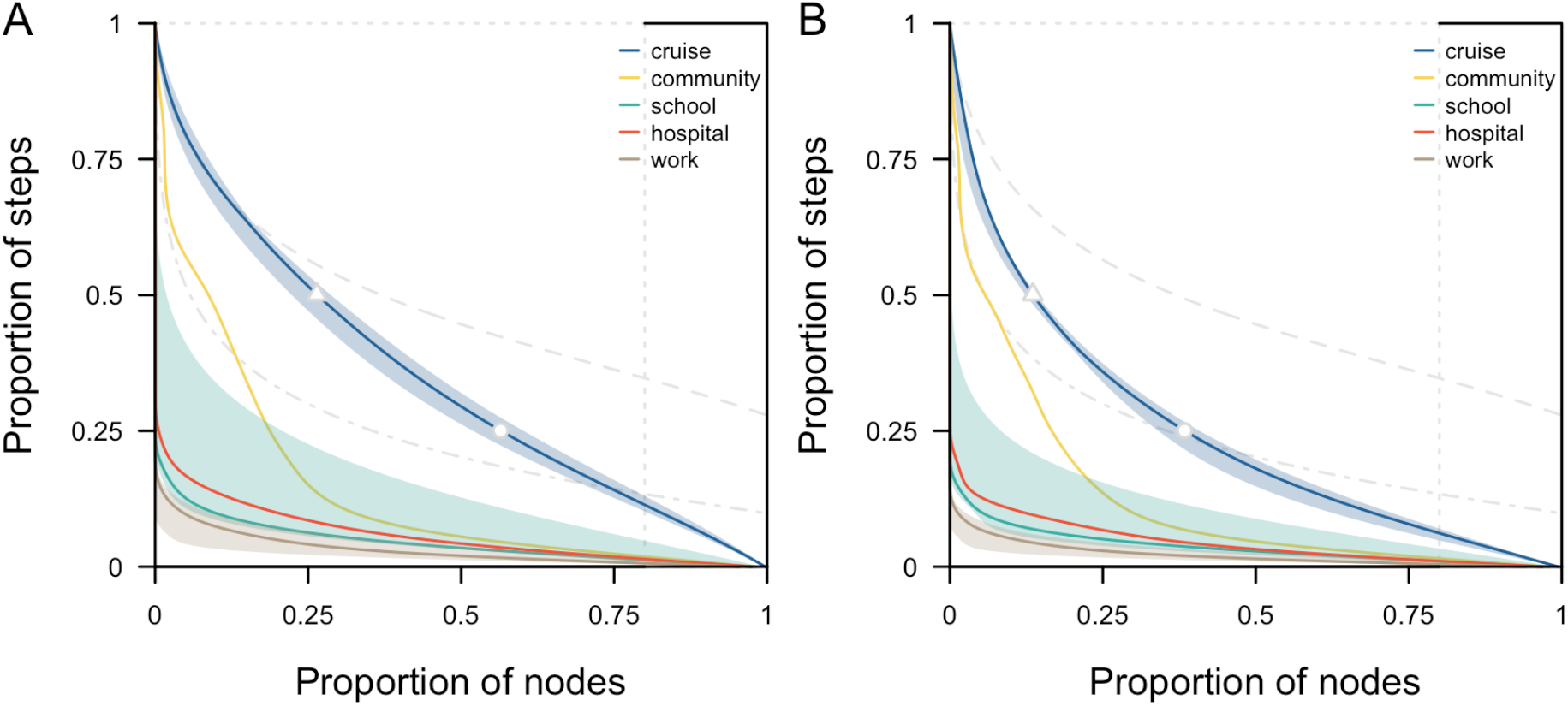
Proportion of superspreaders and superspreading events in respective networks, estimated based on (A) contact episodes or (B) contact duration. For reference, grey lines represent homogeneous static network (dotted), homogeneous dynamic network in 25 timesteps (dashed) and overdispersed dynamic network in 10 timesteps (dot dashed). Cutoff marks for the proportion of individuals in the cruise networks who were highly connected for more than half the total number of time steps (triangle) and those who were highly connected for less than a quarter of the time (dot) as shown.

### Epidemiological metrics

Although a longer study duration will in theory increase the probability of observing a repeated contact over multiple days, there was some agreement across different networks on the proportion of total measured contacts that occurred in one day out of all days in respective network studies. For studies conducted over three days, the proportion of total contacts that occurred over one-day was 86% (range 83–87%) in the cruises and 82% in the community (Figure 3A). For studies conducted over longer durations of up 10 days of recorded contacts, the proportion of total contacts recorded in a given day was 57% (range 52–60%) in the high schools, 51% in the hospital and 47% (range 38–55%) in the workplace networks (Figure 3A). Across all the networks, over 75% of the contacts either occurred over one-day only or were repeated for less than half the study duration (Figure 3A).

When planning outbreak control measures such as contact tracing, we need to consider the number of unique contacts made per infected individual. If we did not account for repeated contacts over the days and instead assumed the measured number of daily contacts would be made independently each day, we could overestimate the number of unique contacts. With the exception of the community network, we found that we would overestimate the unique contacts by 13–35% across all networks after three days of observation under this independence assumption (Figure 3B). For longer study duration in the schools, this difference between the total and unique contacts was 71% (IQR 35%–110%) after seven days; for workplaces, the difference rose to 73% (IQR 33%–130%) after ten days (Figure 3B).

### Extent of superspreaders and superspreading events

Depending on the level of overdispersion of individual-level contacts in a network and the duration of observation, our ability to correctly predict highly connected individuals in a given time period will vary. For a homogenous static network, 80% of the population accounts for 80% of the contacts made. As such, 80% of the population would be identified as highly connected across all the time steps while the remaining 20% of the population would never be identified in this group (Figure 4, dotted lines). For a fully dynamic homogeneous network with 25 time steps, 80% of the population accounts for 80% of the contacts in each time step. Given changes in the network structure over the time steps, only 40% of the population would be identified for at least half the total number of time steps. For a fully dynamic overdispersed network with 10 time steps, 50% of the population accounts for 80% of the contacts in each timestep. Consequently, only 5% of the population would be identified in at least half the observations. We found that as networks transition from homogeneous to overdispersed, and as the duration of observation increases, the proportion of highly connected individuals that can be identified consistently is reduced.

Real-world networks with higher levels of contact retention had a higher probability of correctly predicting frequent, highly connected individuals but these individuals only accounted for less than 30% of the population. These are individuals who account for the top 80% of the contact episodes for at least half of the number of observed time steps (i.e. potential superspreaders, top left region of each panel in Figure 4). In real-world cruise contact networks, 26% (range 22%– 29%) of the population were predicted to fall into this ‘potential superspreader’ category. The remaining population are individuals who have high connections but for short periods of time only. These are individuals who are likely to drive superspreading events (i.e. bottom right region of each panel in Figure 4). In particular, 44% (range 40%–48%) of the population were identified for less than a quarter of the observed time steps (Figure 4A). In the community network, 9% of the population would be predicted to be potential superspreaders while 81% of the population are likely to drive superspreading events for less than a quarter of the time (Figure 4A). The proportion of the population identified as potential superspreaders was less than 5% in the high school, hospital and workplace networks; the majority of the individuals would, if anything, drive superspreading events instead (Figure 4A). Similar trends were observed when analysing the proportion of the population that accounted for the top 80% of the contact duration (Figure 4B).

## Discussion

Using real-world contact data collected from a variety of settings over different days and population sizes, we assessed the key structural properties of temporal networks that drive transmission processes and, hence, influence the effectiveness of outbreak control measures. We estimated that most individuals in each social context had high levels of connectivity with others for less than a quarter of the study duration. Contact retention and the type of contacts driving this retention varied across settings, emphasising the need for tailored outbreak analysis and control strategies for different settings.

In our analysis, we compared the properties of the real-world temporal networks relative to static and fully dynamic networks, normalised by the population size. This enabled us to contextualise our findings and allow for appropriate comparison across different networks. In particular, our study highlighted an inherent difficulty in predicting superspreaders over time across different settings (6). In cruise data, the high level of consistency in identifying highly connected individuals (i.e. 26% of the population identified to account for the top 80% of the contacts in more than half the total observed time steps) was likely influenced by the prevailing COVID-19 restrictions onboard during the study. Passengers and crews were encouraged to remain within their travel or working groups and to practise physical distancing from other groups (11). However, the level of consistency in identifying highly connected individuals was generally low in all other networks. More than 80% of the population was identified to be highly connected for only a short period of the study duration. Targeting small groups of infectious individuals with high levels of connectivity has been shown to, in theory, produce an effective and efficient reduction in transmission, but such studies were largely based on static networks (23, 24). In contrast, our study showed that if we were to sample a network for a few days or a short period of time, and target individuals with high measured connectivity, this level of connectivity would generally turn out to be much lower if data collection were to be repeated in the near future. As such, when designing interventions to identify potential ‘superspreaders’, we would need to target a greater number of individuals than basic theory from static networks suggests in order to achieve the same reduction in transmission.

When an outbreak occurs, outbreak control policies often target subpopulations rather than individuals given the lack of information on contact patterns (15). Across most social settings we analysed, contacts between individuals in the same social group (e.g. same cabin, department or school class) dominated interactions, even if retention of these contacts was variable. For high schools and workplaces, we estimated low contact retention even when most of these contacts were formed between individuals of the same class. This result corroborates previous findings indicating low levels of repeated contact among household contacts for those residing in dormitories (14).

When implementing outbreak control policies, our results suggest it is important to consider if the priority is to reduce disease introductions, or reduce transmission if introduced to a locality, and thus, which is the appropriate individuals or subpopulations to target with restrictions. In schools and workplaces, the majority of close contacts were from individuals of the same department or class, implying that targeted rather than school- or workplace-wide closures could still help to minimise disruption to activities. This would be particularly relevant if disease prevalence in the wider population is low and the likelihood of introductions to other departments or classes is low. In contrast, for settings such as hospitals, contacts from both the same (e.g. nurse-nurse contacts) or different (e.g. patient-nurse contacts) departments are likely to be retained over consecutive time steps. This higher proportion of contacts between different departments is expected given the multi-faceted roles of healthcare workers (18). Thus, more stringent measures to reduce the risk of nosocomial outbreaks starting is highly important to avoid disruptions to hospital functions.

While the use of detailed contact data to plan quarantine measures can provide an upper limit on the resources required (7, 9), our results suggest the occurrence of repeated contacts would mean that simple analysis, based on cross-sectional data collection that assumes independence of contacts, would generally overestimate the resources required for contact tracing each case. With the occurrence of pre-symptomatic transmission for SARS-CoV-2 (25, 26) and delays from symptoms onset to testing to isolation (27, 28), contact tracing would involve the identification of cases over 3–11 days and repeated contacts arising from regular daily activities would imply that the actual contacts made over this period are 20–70% lower than the sum of all the contact episodes recorded independently on each day.

There are some limitations to our study. First, we focused on the network and epidemiological metrics between pairs of contacts. We did not study the changes in clustering on temporal networks and overlay the dynamics of infectiousness profiles on these networks. As such, this limits our ability to make conclusions on the impact of temporal contacts on outbreak size, time to outbreak extinction and herd immunity thresholds. Nevertheless, the current study is a first step in characterising temporal networks. Our ‘retention index’, 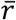, quantifies the retention of contacts in temporal networks relative to static and highly dynamic networks. Furthermore, we analysed the type of contact pairs that are likely to be retained and highlighted the implications to control measures. Future studies could extend this metric to account for higher-order network properties. This would allow us to better understand the impact of time-varying contacts on disease transmission and study the feasibility of using simpler static networks or compartmental models.

Second, different devices were used to measure the networks in different studies. They could either detect face-to-face interactions or RFID signals from all directions. As each device has a different calibration, the measured differences between the networks can be an outcome of the data collection process or due to inherent differences in the context setting. As such, in the main analysis, we defined the contact duration and delay between contacts based on the characteristics of each network (Table 1). In our sensitivity analysis, we standardise the duration and delay. The changes in 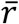 for different networks were similar in both analyses. Hence, the impact of the device setting on the overall observed contact patterns was not expected to be significant. Thirdly, real-life contact typically exists in an open population and thus not every contact was captured in these network studies. If these missed contacts were to occur in specific sub-populations this may result in a shift in the proportion of retained contact types. Furthermore, the level of connectivity in missed contacts is unknown. As such, our analysis could over- or underestimate the proportion of superspreaders and superspreading events. However, our findings would remain valid if we assume that the missingness is independent of the level of connectivity and can occur in any subpopulation.

Our analysis highlights the difficulty in identifying highly connected individuals unless real-world contacts are surveyed at high resolution over several days. However, we did find more consistency in contact patterns among specific settings and social groups. Hence, outbreak control measures that target key settings or at-risk subpopulations are likely to be more effective than targeting specific individuals if currently available data approaches continue to be used. Comparing the dynamics of such real-world temporal networks and corresponding outbreak data would further advance our understanding of the risk of different contacts in practice.

## Ethics

The study was approved by the London School of Hygiene & Tropical Medicine Observational Research Ethics Committee (ref. 25727). All data and analysis were collected and performed in line with the Infectious Diseases Act in Singapore which permits the collection and publication of surveillance data.

## Data availability

Data and code available at https://github.com/rachaelpung/temporal_networks

## Funding Additional Information

RP acknowledges funding from the Singapore Ministry of Health. J.A.F. was supported by funding from BBSRC (BB/S009752/1) and NERC (NE/S010335/1 and NE/V013483/1). AJK was supported by a Sir Henry Dale Fellowship jointly funded by the Wellcome Trust and the Royal Society (grant 206250/Z/17/Z). The funders had no role in study design, data collection and interpretation, or the decision to submit the work for publication.

## Author Contributions

Conceptualization: RP and AJK. Methodology: RP, JAF, TWR, TR and AJK. Investigation: RP, JAF and AJK. Visualization: RP, JAF, TWR and AJK. Supervision: AJK. Writing, original draft: RP. Writing, review & editing: all authors. All authors read and approved the final manuscript.

