## Supplementary Figure for "Temporal contact patterns and the implications for predicting superspreaders and planning of targeted outbreak control"

**Supplementary Figure 1** Changes in contact retention index, $\bar{r}$, over time for (A-D) four cruise networks, (E) one community network, (F-H) three high school networks, (I) one hospital network, (J-K) two workplace networks. The duration of observation for each day is not necessarily the same across all studies.

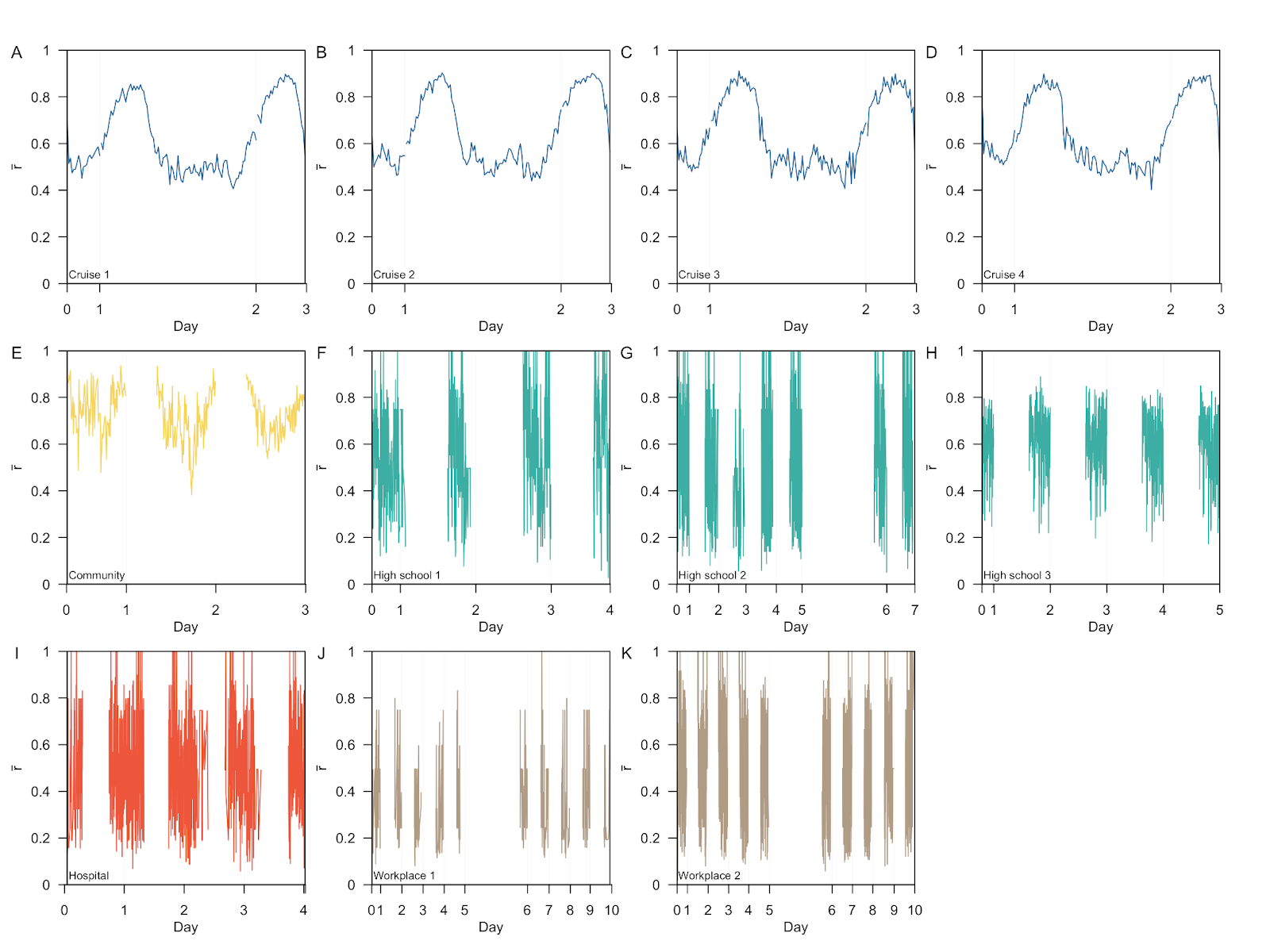

**Supplementary Figure 2** Contacts patterns in different settings for contacts assuming undirected contacts in all networks, (a) distribution of contact retention index, $\bar{r}$, over consecutive timesteps, (b) proportion of each type of contact retained for respective $\bar{r}$

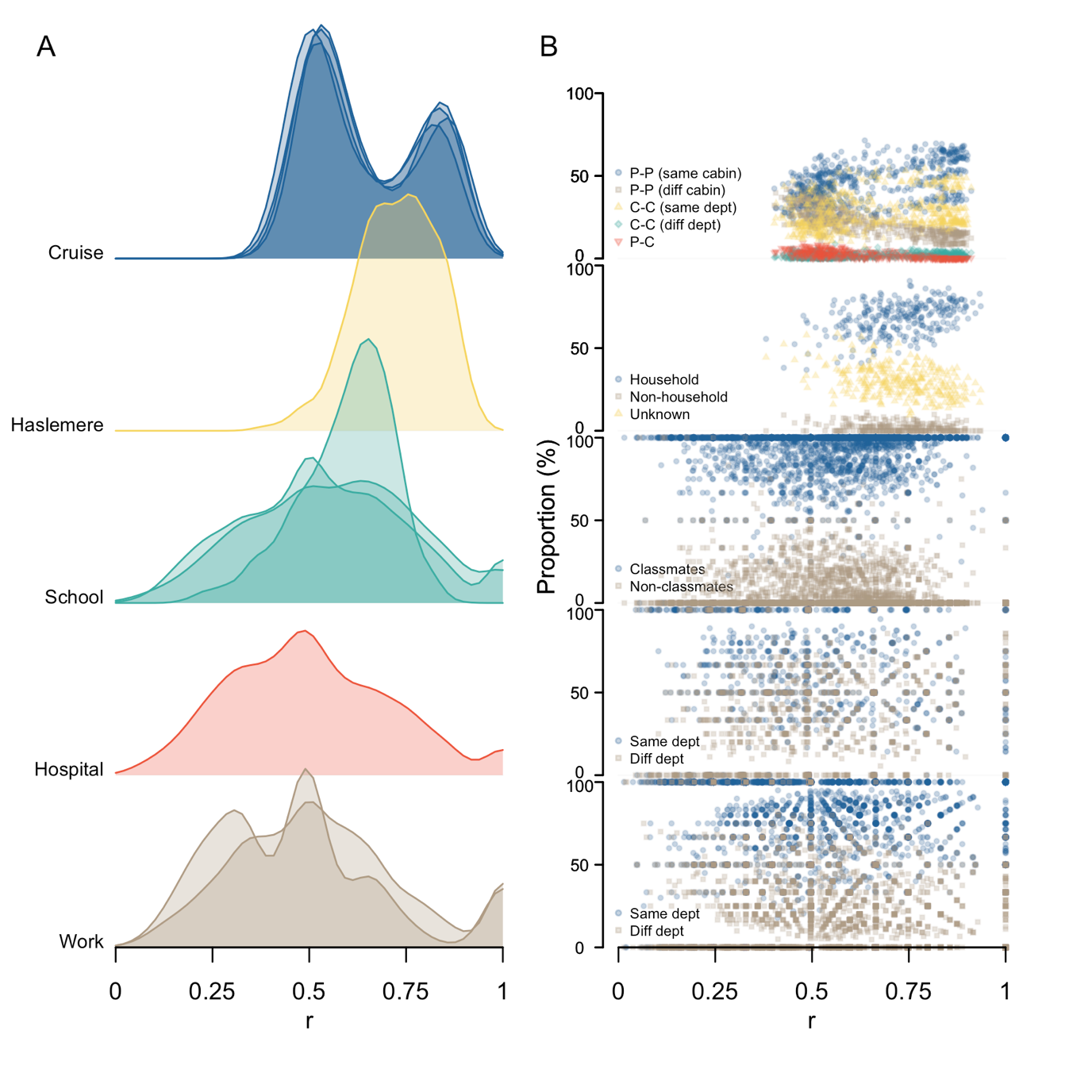

**Supplementary Figure 3** Contacts patterns in different settings for contacts formed in a fixed time window of 1-hr, (a) distribution of contact repetition, $\bar{r}$, over consecutive timesteps, (b) proportion of each type of contact retained for respective $\bar{r}$

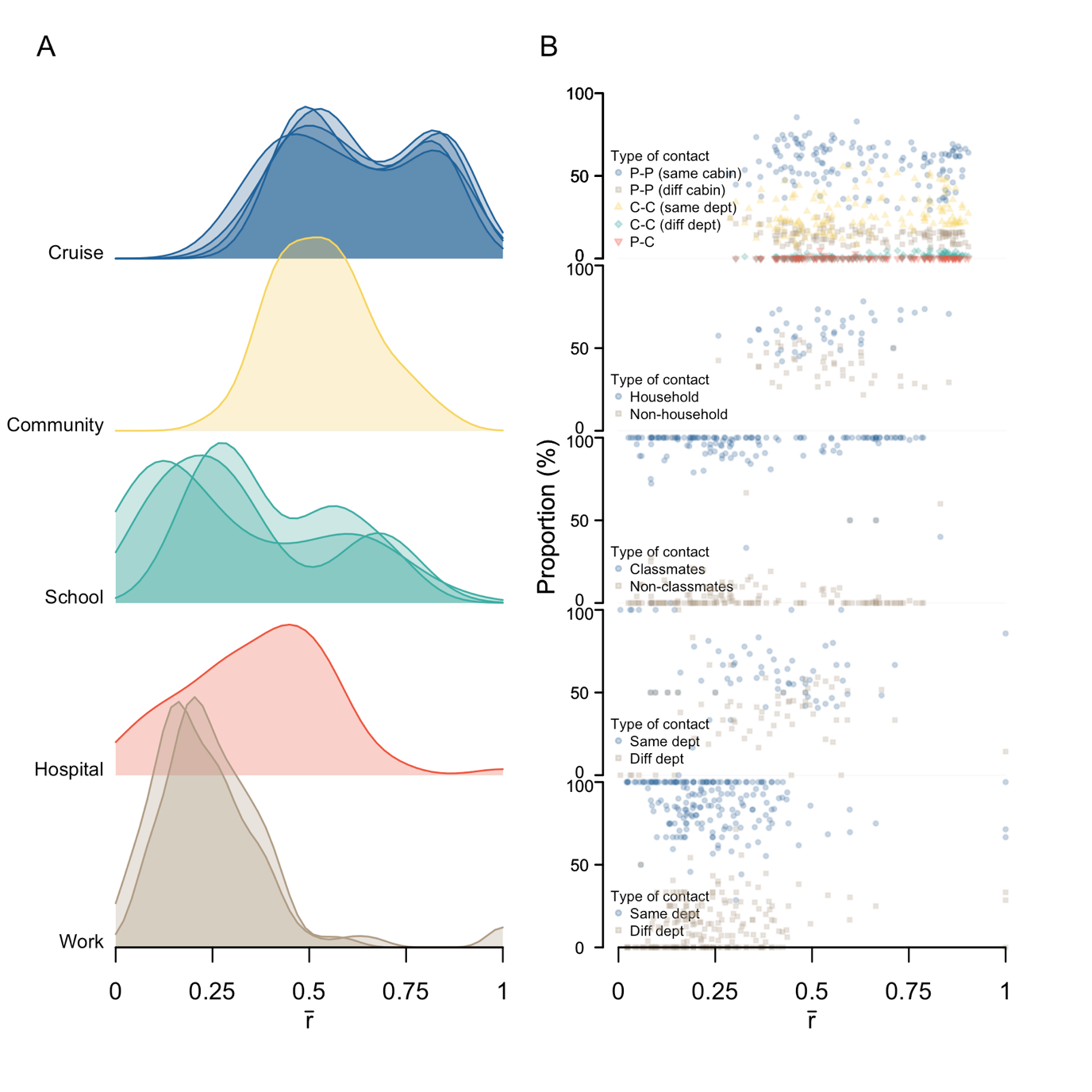

**Supplementary Figure 4** Contacts patterns in different settings for contacts formed in a fixed time window of 15-min, (a) distribution of contact repetition, $\bar{r}$, over consecutive timesteps, (b) proportion of each type of contact retained for respective $\bar{r}$

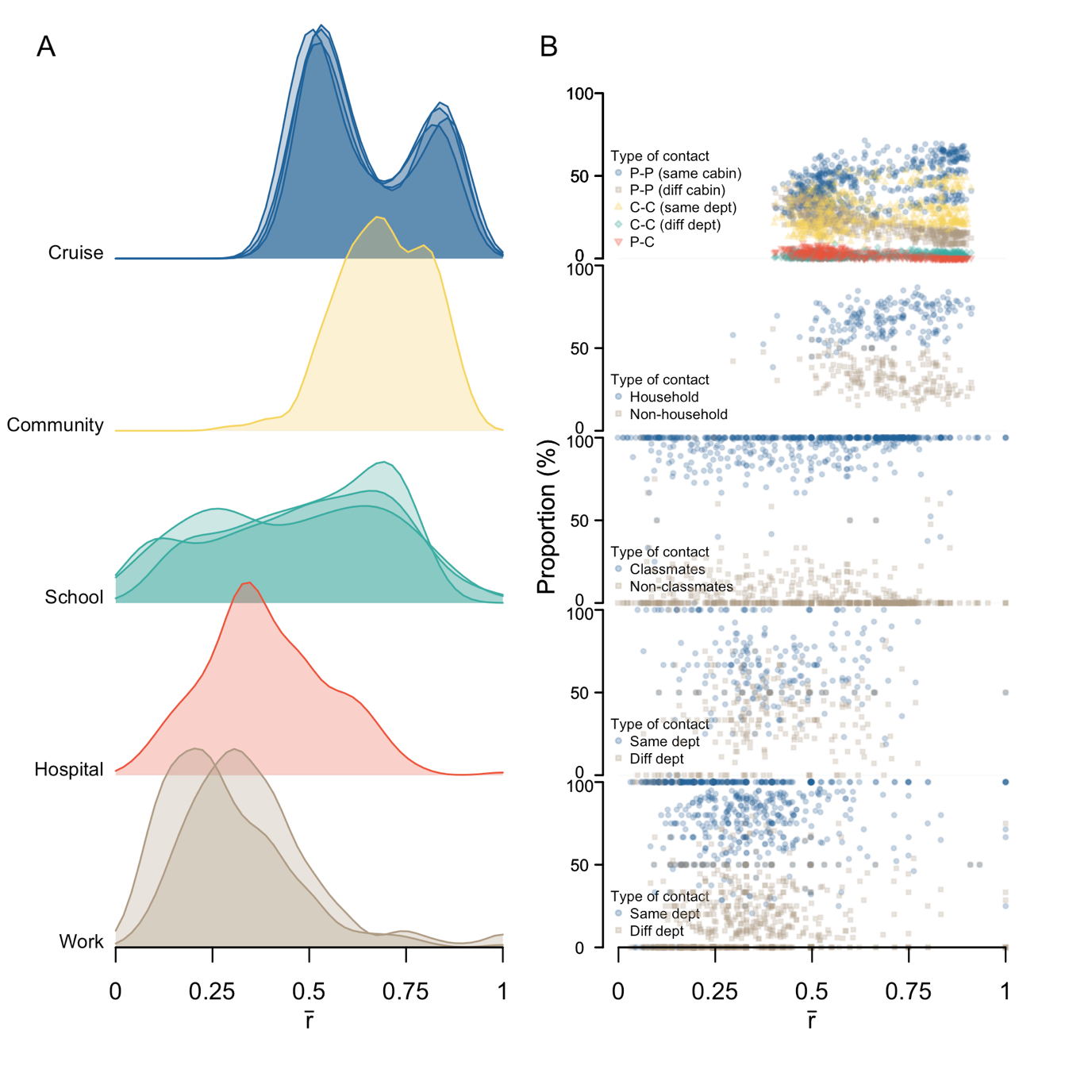

**Supplementary Figure 5** Proportion of ‘superspreaders’ and ‘superspreading events’ in respective networks (coloured). Individuals that account for the top 80% of the contact (A) episodes or (B) duration in a day were identified.

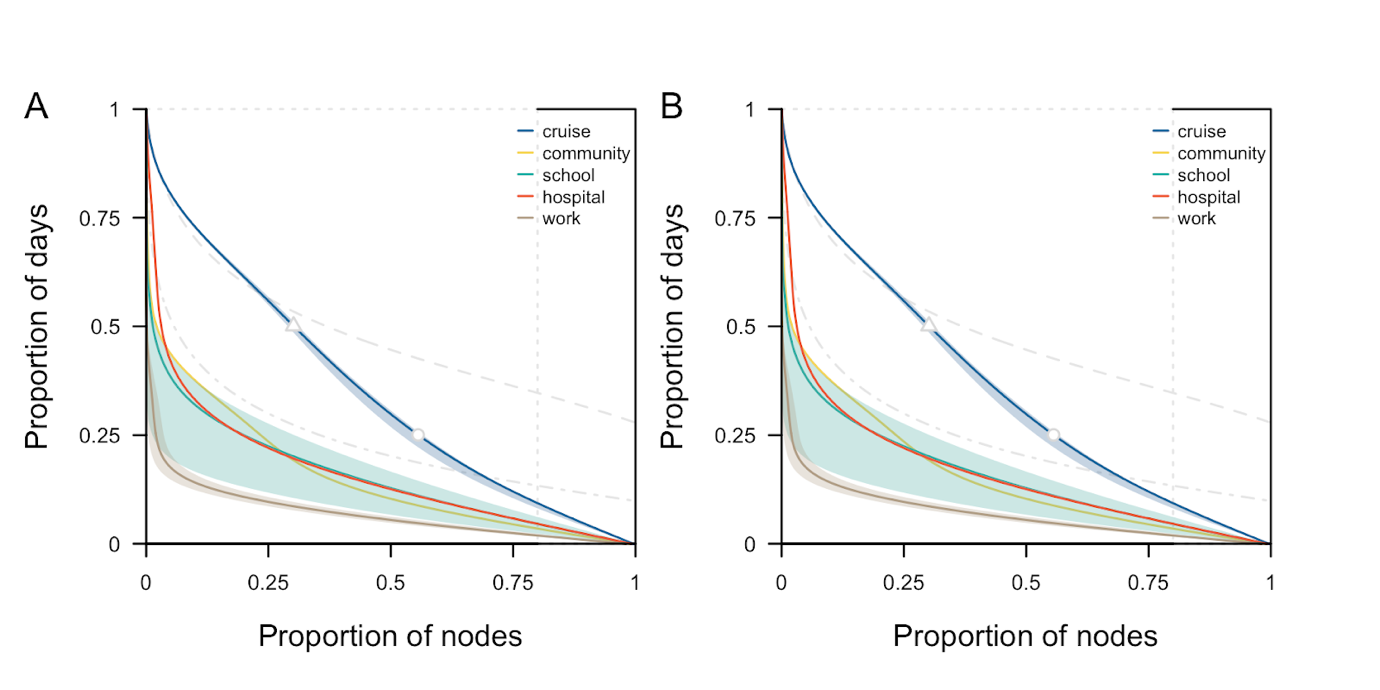
